# Clinical Predictors of 3-Months Isoniazid Rifapentine (3HP) - Related Adverse Drug Reactions (ADR) During Tuberculosis Preventive Therapy. (PAnDoRA-3HP study): An Observational Study Protocol

**DOI:** 10.1101/2024.06.01.24308310

**Authors:** Christine Sekaggya-Wiltshire, Irene Mbabazi, Ruth Nabisere-Arinaitwe, Grace Banturaki, Lucy Alinaitwe, Brian Otalo, Florence Aber, Juliet Nampala, Rogers Owor, Josephine Bayiga, Eva Laker Agnes Odongpiny, Barbara Castelnuovo, Jonathan Mayito, Moorine Sekadde, Jotam G. Pasipanodya, Stavia Turyahabwe, Stella Zawedde-Muyanja

## Abstract

**Introduction:** Tuberculosis (TB) is the leading infectious cause of death globally. Despite WHO recommendations for Tuberculosis Preventive Therapy (TPT), challenges persist, including incompletion of treatment and adverse drug reactions (ADRs). There is limited data on the 3-month isoniazid and rifapentine (3HP) pharmacokinetics, pharmacogenomics and their relation with ADRs. Our study aims to describe the pharmacokinetic and pharmacogenomics of 3HP used for TPT, the ADRs and their association with completion rates, and TPT outcomes, providing vital insights for TB control strategies in resource-limited settings.

**Methods:** This is an observational cohort study with a nested case-control study. We enrolled consecutive patients initiated on TPT using the 3HP regimen. These are followed up bi-weekly and then monthly during the active phase of treatment and 3 monthly for 2 years following completion of TPT. ADR evaluation includes clinical assessment and liver function tests. Cases are selected from those who experience ADRs, and controls from those who do not. Serum isoniazid and rifapentine concentrations are measured and pharmacogenomic analysis for NAT2 and CYP2E1 polymorphisms are done. Participants are followed up for 2 years to determine TPT outcomes.

**Analysis:** The safety profile of 3HP will be assessed using descriptive statistics, including proportions of patients experiencing ADRs and grade 3 or above events related to treatment. Chi-square tests and regression models will determine predictors of ADRs and their impact on treatment completion. Pharmacokinetic-pharmacodynamic modeling will establish population parameters and factors influencing rifapentine and isoniazid concentrations.

## INTRODUCTION

23% of the people who developed Tuberculosis (TB) in 2022 were from the WHO Africa region (1). In 2019, Africa recorded 2.46 million new tuberculosis (TB) cases, a quarter of global cases, with the highest incidence rate worldwide of 231 cases per 100,000 (2). HIV-TB rates are 5 times higher in Africa than the worldwide average. The incident rate of TB in Uganda is among the highest in the world at 200 cases per 100,000 and like the rest of Africa, the rates of HIV-TB are one of the highest (81 cases per 100,000) [1].

TB’s global burden extends beyond active cases, with a quarter of the global population harboring latent TB infection (LTBI) (3). Approximately 5%–10% of persons with LTBI progress to active TB disease during their lifetime (4-6), however, Tuberculosis Preventive Therapy (TPT) reduces TB incidence by 40% (7, 8), which is crucial in order to achieve WHO’s goal of ending TB by 2030 (9). WHO recommends TPT for all persons-living-with-HIV (PLWH) and TB contacts with latent TB. (4). Uganda recently rolled out TPT using the three months of weekly isoniazid and rifapentine (3HP) in 2022 (10). 3HP has better completion rates, and is non-inferior to the 6 months of isoniazid (11-15). However, there is limited data on 3HP safety and drug pharmacokinetics in routine clinical settings from Africa, especially in the pediatric and adolescent population (13).

Several studies have demonstrated pharmacokinetic variability of isoniazid and that of rifamycins in people receiving treatment for active TB disease (16-18). In a recent study evaluating the pharmacokinetics of isoniazid in 30 people living with HIV, high isoniazid exposures were found, however the ADRs were not linked to the 3HP. Polymorphisms in NAT-2 is associated with variability in isoniazid concentrations and may contribute to the occurrence of ADRs (19). In Taiwan, 22.8% of participants were slow metabolizers of isoniazid and NAT2 rs1041983 polymorphisms were associated with severe ADRs (20). The mechanism underlying some ADRs including isoniazid-induced hepatotoxicity has been referred to as a ‘metabolic idiosyncrasy’, because it is incompletely understood (21, 22). The reasons for inter-individual variability of isoniazid concentrations, especially in LTBI patients, is still poorly described, and how those variable exposures might contribute to ADRs is still unknown (16, 21-24). Isoniazid and its metabolites may be directly cytotoxic, particularly in the presence of rifampin or similar drugs that induce cytochrome P450. Other molecular mechanisms might contribute to ADRs including immune-mediated processes influenced by HLA immunogenetic markers (25). The effect of recently described HLA genotypes and other drug metabolizing enzymes, such as cytochrome 2E1 (CYP2E1), and Uridine 5’-diphospho-glucuronosyltransferase (UGT), on relative concentrations of isoniazid and rifamycins hepatotoxicity risk in slow versus fast acetylators needs to be elucidated further.

Majority of the data available on 3HP is from clinical trials which we may not be able to entirely extrapolate to the “real life” settings. This study will generate and compare pharmacokinetic, pharmacogenomic and immunogenetic data from patients in a routine clinical setting experiencing ADR and matched controls without ADR to determine the mechanism underlying inter-individual variability of ADR in LTBI patient populations. We therefore aimed to describe the safety profile of 3HP among people receiving tuberculosis preventive therapy and the effect of ADRs on TPT completion rates. We also sought to describe the pharmacokinetic and pharmacogenomic determinants of the ADRs. We hypothesize that 3HP related ADR will be minimal and those who do experience ADRs are more likely to discontinue treatment.

## Methods and Analysis

### Study Site

The study is being conducted at the Infectious Diseases Clinic (IDI) at Mulago National Referral Hospital, Kampala, Uganda, where over 8000 HIV infected patients are managed annually. The study is also taking place in Kampala City Council Authority (KCCA) health facilities.

### Study Population

Patients are enrolled if they meet the following inclusion criteria; 1) individuals of any age who have been initiated on TPT using the isoniazid/rifapentine regimen according to standard of care, 2) both PLWHIV and HIV-uninfected individuals are eligible and 3) subjects who are willing and are able to comply with study procedures.

Patients are excluded from the study if they meet any one of the following criteria; 1) women who are pregnant at the time of screening and 2) individuals who have already received 3HP for more than one month

### Study Design

This is a cohort study including 651 participants with a nested case control study (150 cases and 150 controls). Over 18 months, we enrolled consecutive patients who are already initiated on TPT by the facility clinician as per standard of care. At enrolment, baseline clinical and demographic information are collected including: age, sex, HIV status, type of ART where applicable, weight/BMI, concomitant medications and comorbidities, prior history of TB treatment, and history of substance abuse. Blood samples for baseline Alanine aminotransferase (ALT) and total bilirubin are also drawn.

The 3HP is provided through standard of care and administered to the patient as a weekly dose of rifapentine and isoniazid for up to 12 doses (3 months). Treatment is self-administered at home except on the study visit where treatment will be given as directly observed therapy (DOT) and in order to conduct the pharmacokinetic analysis. We assess adherence by self-report and review of the TPT card. Number of doses missed are recorded.

### Nested Case Control Study

Cases are selected from patients within the cohort study who have developed adverse events that are related to the drug based on the Naranjo score. Controls are selected by incidence density sampling from those within the cohort study who do not develop any ADR related to drug. Cases and controls are matched by duration on 3HP, sex, and age.

### Follow up

All enrolled participants are followed to evaluate for ADRs after 2 weeks from enrolment and then monthly during the 3 months of 3HP treatment. Following completion of 3HP, 3 monthly visits are conducted for 2 years.

### Primary outcomes

Our primary outcomes are adverse drug reactions and TPT completion.

#### 1) Adverse drug reactions

Adverse drug reactions are assessed using clinical history, physical examination and laboratory tests (ALT and total bilirubin). The ALT and bilirubin measurements are performed at the IDI Core laboratory, which is a College of American Pathologists certified laboratory.

Participants who present with symptoms suspected to be ADRs are further evaluated and additional laboratory tests may be performed. ADRs that may have been experienced between visits are also evaluated and recorded. ADRs are assessed for relationship to the drug using the Naranjo scale (26). Follow-up is conducted to determine the outcome of the ADR and whether subsequent doses of TPT are taken or not. Division of AIDS (DAIDS) version 2.2 (27) is used to grade the severity of ADRs.

#### 2) TPT completion rates

TPT completion is defined as taking all 12 doses of HP within 16 weeks of starting treatment. For those who undergo interruption of treatment for any reason, the total duration of their TPT is recorded. Interruption and re-starting of TPT is in accordance to national treatment guidelines (10).

### Secondary outcome

Our secondary outcome is efficacy of 3HP in terms of TB reactivation or a TB episode following TPT within 2 years of treatment completion

#### Evaluation for TB after TPT

All participants in the cohort are followed up for 2 years following 3HP initiation. Screening for signs and symptoms of TB is conducted at the monthly and 3-monthly visits using the intensified case finding form. A genexpert, chest X-ray or urine LAM is requested in those who present with presumptive TB.

### Pharmacokinetic analysis

Blood sampling for PK analysis is done for patients (cases and controls) within the nested case-control study to measure rifapentine and isoniazid concentrations. The first PK sampling occurs within 2 weeks of being selected as a case or control. The second PK sampling is performed before or at the month 3 visit. A meal is provided prior to drug intake. Blood samples are drawn pre-dose, at 4hrs and 24 hours post-dose. Time and date of last dose is noted on these occasions.

Drug concentrations are measured using ultra-performance liquid chromatographic-tandem mass spectrometry or high-performance liquid chromatography.

### Pharmacogenomic analysis

Blood sampling is performed for polymorphisms in CYP2E1 (rs2070673). In addition, NAT2 acetylator status is determined using a three-single nucleotide polymorphism panel of 191G > A (rs1801279), 341T > C (rs1041983) and 857G > A (rs1799931).

DNA extraction is performed using commercially available kits according to the manufacturer’s instructions. Genotyping is conducted using real-time PCR allelic discrimination using the TaqMan assay (Applied Biosystems, Warrington, U.K.) at the IDI Translational laboratory.

HLA genotyping for class 1 will be performed using polymerase chain reaction (PCR). Allele-specific PCR, using sequence-specific primers will be performed according to the protocol and recommendations of the manufacturer.

**Table 1:**
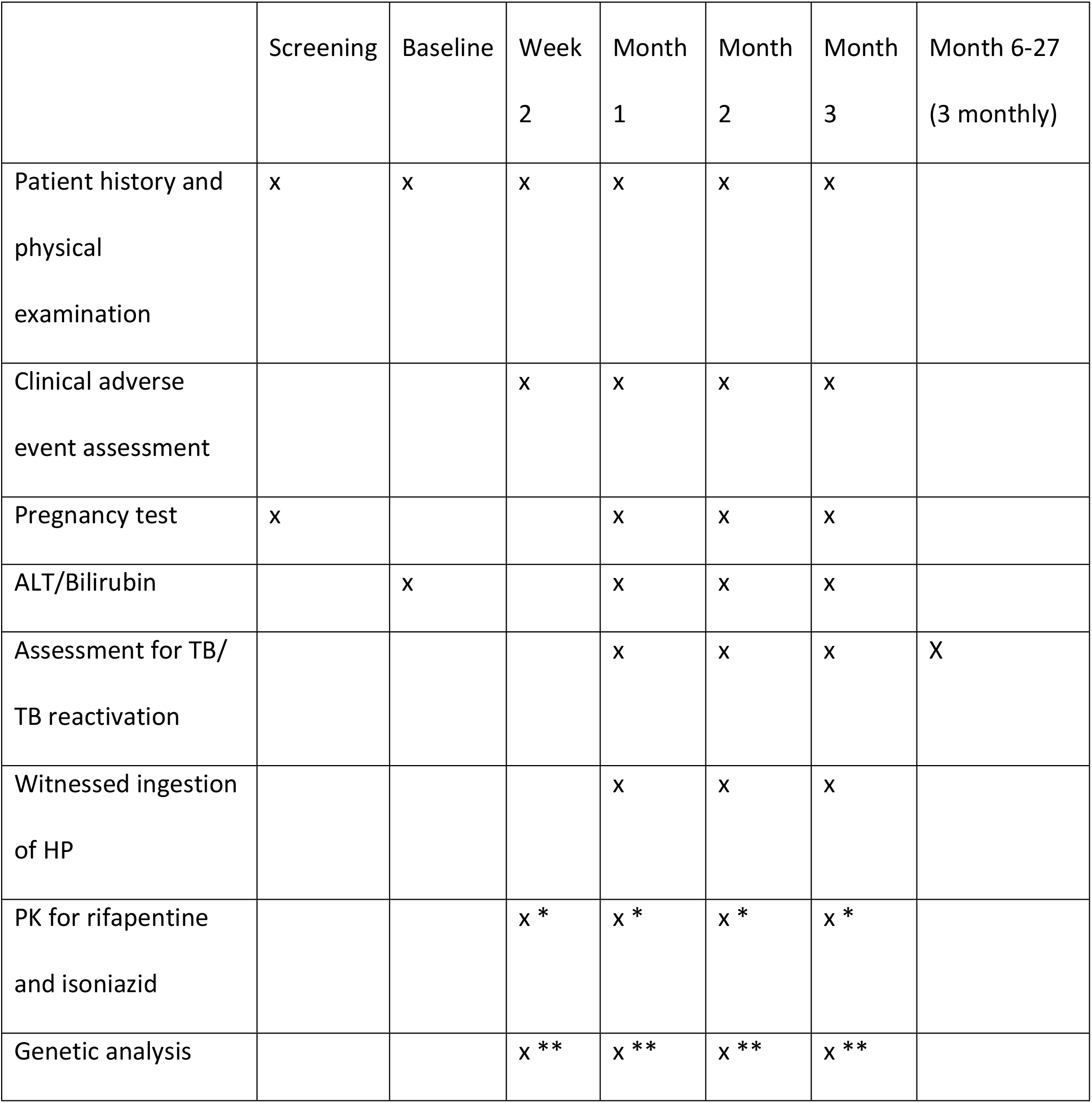
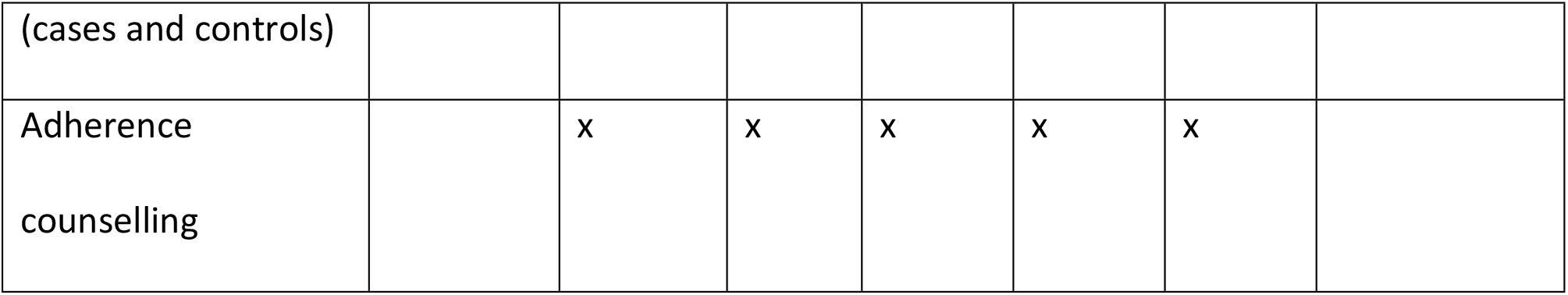
Study procedures.

### Patient and Public Involvement Statement

In the exploration of adverse events associated with 3HP within Uganda’s routine clinical setting, the Patient and Public Involvement (PPI) section stands as a testament to our dedication to inclusivity and collaboration. At the outset, patients and the public were intimately involved in shaping the study’s direction, with their insights guiding the formulation of research questions. Drawing from their lived experiences, preferences, and priorities, our study aims to resonate deeply within the community it serves. Throughout the design and execution phases, patients and the public have been indispensable partners. Their input has steered decisions concerning study design, the selection of outcome measures, and the crafting of recruitment strategies.

Furthermore, their ongoing engagement extends to critical decisions regarding the plans for disseminating study findings. By fostering this level of meaningful engagement, our study not only amplifies the voices of those affected by tuberculosis but also ensures that our findings are both accessible and actionable within Uganda’s healthcare landscape, contributing to more effective TB control strategies in resource-limited settings.

### Statistical Analysis

Safety profile of 3HP will be described using descriptive statistics including proportions. We will determine the proportion of patients who had any ADR and those with grade 3 and above events with their relatedness to the treatment. Chi-Square tests and regression models will be used to determine predictors of ADR and determine effect of ADRs on TPT completion. We will determine the proportion of patients who experience TB reactivation over 2 years stratified by risk category. Time of follow-up will be calculated as person-years and time to reactivation will be determined. This analysis will be performed using STATA 14.0.

#### Pharmacokinetic-Pharmacodynamic modelling

Non-linear mixed effects modelling technique will be used develop a model to establish the population parameters for rifapentine and isoniazid and the variability around these parameters. Using this approach, we will then determine the PK parameters: area under the concentration-time curve (AUC) and maximum concentrations (Cmax) of rifapentine and isoniazid and the factors that affect these PK parameters including age, sex, and pharmacogenomic variants in NAT-2 and CYP2E1.

### Ethics

Ethical approval for this study was sought and approvals received from the Infectious Diseases Institute Research and Ethics Committee (IDI-REC), IREC Ref: 001/2021 and the Uganda National Council of Science and Technology (UNCST), UNCST Folio Number: HS1582ES.

Written Informed consent is obtained from all participants. Children between 8 and 17 years are requested for written assent, in addition to the written consent from their parents or legally authorized representative. For children less than 8 years, written consent is sought from their parents/ legally authorized representative. The study is conducted in accordance with good clinical practice and declaration of Helsinki.

### Study Status

The study began recruitment of study participants on 24^th^ February 2022 and completed recruitment on 4^th^ August 2023. All participants have completed TPT and are undergoing follow up for TB reactivation. Follow-up will be completed in September 2025.

## Discussion

### Strengths and Limitations of this study

- Comprehensive Scope and design: The study encompasses multiple healthcare facilities in Uganda, ensuring diverse patient populations.. The long-term follow-up over 2 years post-TPT completion allows for robust evaluation of outcomes and adverse events.
- The nested case-control study will allow us to critically evaluate population pharmacokinetics of those who had adverse events. Currently there is limited data that describes the pharmacokinetics of once weekly isoniazid and rifapentine. Incorporating genetic analysis enhances understanding of pharmacokinetic data and individual responses to treatment.
- A limitation of this study is that Directly observed therapy (DOT) is not performed since this study is being conducted in a programmatic setting. This can lead to reporting bias. However, DOT is performed during the monthly visits and visits where pharmacokinetic analysis is to be performed.

### Dissemination

De-identified genotype data from 300 patients will be shared after publication. The protocol and phenotype data will be publicly accessible. Abstracts will be submitted to conferences, and a manuscript will be published post-study.

### Participant Withdrawal

Subjects may withdraw from the study at any time at their own request, or they may be withdrawn at any time at the discretion of the investigator or sponsor for safety or behavioral reasons, or the inability of the participant to comply with the protocol required schedule of study visits or procedures. Participants who are withdrawn from the study can be replaced.

If a participant does not return for a scheduled visit, every effort will be made to contact the participant. In any circumstance, every effort will be made to document participant outcome, if possible. The investigator will inquire about the reason for withdrawal and follow-up with the participant regarding any unresolved adverse events (AEs).

If the participant withdraws from the study, and also withdraws consent for disclosure of future information, no further evaluations will be performed, and no additional data will be collected. The study team may retain and continue to use any data collected before such withdrawal of consent.

## Data Availability

No datasets were generated or analysed during the current study. All relevant data from this study will be made available upon study completion.

## Author contributions

This study was conceptualized by CSW and SZM. The study design was conceived by CSW, SZM, EL, JM, JP, BC, ST, MS. IM, RN, LA, FA, BO, RO, JB, JN assist with data collection while JP and GB assist with data analysis. IM, RN and CSW wrote the manuscript. All named authors adhere to the authorship guidelines of Trials. All authors have agreed to publication.

## Notes

**Funding statement:** The National Institute of Allergy And Infectious Diseases of the National Institutes of Health under Award Number R01AI160434 supported research reported in this publication. The content is solely the responsibility of the authors and does not necessarily represent the official views of the National Institutes of Health

### Competing Interest Statement

The authors have declared no competing interest.

### Funding Statement

Yes

### Author Declarations

Ethical approval for this study was sought and approvals received from the Infectious Diseases Institute Research and Ethics Committee (IDI-REC), IREC Ref: 001/2021 and the Uganda National Council of Science and Technology (UNCST), UNCST Folio Number: HS1582ES. Written informed consent is obtained from all participants. Children between 8 and 17 years are requested for assent, in addition to consent from their parents or legally authorized representative. For children less than 8 years, consent is sought from their parents/ legally authorized representative. The study is conducted in accordance with good clinical practice and declaration of Helsinki.

